# SARS-CoV-2 specific immune-signature in direct contacts of COVID-19 cases protect them from contracting disease: A Retrospective Study

**DOI:** 10.1101/2021.03.11.21253367

**Authors:** Sunil K. Raghav, Kaushik Sen, Arup Ghosh, Sudeshna Datta, Abdul Ahad, Atimukta Jha, Sanchari Chatterjee, Sandhya Suranjika, Soumya Sengupta, Gargee Bhattacharya, Omprakash Shriwas, Kiran Avula, Jayasingh Kshatri, Punit Prasad, Ajay K. Parida

## Abstract

The response to SARS-CoV-2 is largely impacted by the level of exposure and the status of immunity. The nature of protection shown by direct contacts of COVID-19 positive patients is quite intriguing to note. We aimed to study the immune differences reinforcing contact individuals in circumventing the disease. Our observation showed direct contacts of PCR positive patients developed elevated neutralizing antibody titres and cytokine levels. On the other hand, single cell data revealed differential usage of V(D)J genes and unique BCR clonotypes imparting protective immune signatures.

**Topics:** serologic tests, immunoglobulin a, immunoglobulin g, immunoglobulin m, antibody titre; cytokine levels; virus neutralization; V(D)J sequencing; BCR clonotypes

## Background

Off late the incidence of new cases of COVID-19 has declined remarkably in certain regions which has led to the plausible explanation of hygiene hypothesis or prevalence of antibodies with neutralizing capabilities [1, 2, 3]. The humoral immune response plays a pivotal role in evading viral infections. Several reports have appreciated the role of serum IgA for early neutralizing response against SARS-CoV-2 and their longevity for months after onset of symptoms [4, 5, 6, 7]. The induction of humoral immune response can increase the efficacy of vaccination [8]. Last year during the period of June to July large groups of migrant workers started returning back to their native residence in shared transports. From the SARS-CoV-2 RT-qPCR surveillance information of the migrant workers we observed that some individuals have not contracted the disease even after travelling in close vicinity (proximity <1m) of COVID-19 bearing individuals. To understand the immune status of these individuals (direct contacts <1m proximity) we enrolled three subject groups for our study namely control (CTRL, unexposed to SARS-CoV-2), infected (INF, COVID-19 positive, inception confirmed by RT-qPCR test) and contact (CON, individuals who travelled with SARS-CoV-2 RT-qPCR positive individuals in the same vehicle sitting within ∼1m radius for 3-4 days). We further divided the infected individuals into two subgroups *viz* symptomatic (SYM, showing mild or moderate symptoms) and asymptomatic (ASY) based on disease severity at the time of sample collection. This study has taken into account candidates of both sexes with age ranging from 4 to 60 years.

## Methods

### Enzyme linked immunosorbent assay (ELISA)

For IgA and IgM quantification, RBD (SARS-CoV-2 spike RBD recombinant protein, mFc-Tag, CST # 41701S) antigen at a concentration of 200ng/well was coated in a 96-well high binding micro titre plate (HIMEDIA-EP1) in 1x TBS pH-7.4 for 2h at 37□. Following incubation, the plate was washed 3x times with a wash buffer (TBS containing 0.2% Tween 20). Blocking was done with milk (3% skim milk in TBS containing 0.05% Tween 20) for 1h at 37□. Thereafter wells were washed and 50 µl of serum (1:320 diluted) was added for 1h at 37□. Secondary antibody HRP Goat anti-human IgA (Biolegend #411002) and GtX Hu IgM HRP (Merck Millipore # Lot: 3462097) were used at a dilution of 1:2500 for 1h at 37□ for IgA and IgM quantification respectively. Finally, after washing, 50 µl of TMB substrate (Biolegend # 421101) was added for development of colour for approximately 15 minutes. The reaction was stopped using 2N H_2_SO_4_ and absorbance was measured at 450 nm in a Multiskan reader (Thermo scientific).

For quantification of total COVID-19 and IgG antibodies, kits namely COVID-19 (IgG + Ig M + Ig A) Microlisa and Covid Kawach IgG Microlisa (# IR200196) were used (J. mitra & Co.) respectively. The tests were performed according to the manufacturer’s protocol.

### Bioplex

Human cytokine quantification was performed from serum samples according to manufacturer’s protocol (*Bio-Plex Pro Human Cytokine Screening Panel #*10000092045, Part no-12007283). Briefly, 50 µl of 1x beads were added to the wells and washed 2x times with 200 µl of wash buffer. 50 µl of standards, samples (1:4 diluted) and controls were added and incubated on the shaker at 850 rpm for 30 minutes at RT. Following which in the same manner 25 µl of 1x detection antibody was mixed and incubated. Thereafter, the wells were washed and streptavidin-PE was added for 10 minutes with shaking. Eventually after giving final washes, the samples were resuspended in 125 µl of assay buffer and data acquisition was performed on Bio-Plex 200 System.

### PBMC staining and flow cytometry

Peripheral blood mononuclear cells (PBMCs) were isolated from control, contact and SARS-CoV-2 infected individuals. Cells were washed with RPMI-1640 media containing 10% FBS and proceeded for staining in 1x FACS buffer (3% FBS in 1x PBS). Single cell suspension was counted and blocked with Human TruStain FcX™ Fc Blocking reagent (BioLegend # 422302) for 10 minutes on ice. After blocking, cells were washed with 1x FACS buffer and incubated with conjugated primary antibodies anti-human CD19 PB (eBiosciences # 48-0199-42). Cells from individual patient samples were incubated with anti-human TotalSeq™-C antibodies (C0251 BioLegend # 394661, C0252 BioLegend # 394663 and C0253 BioLegend # 394665), washed and treated with PI viability dye (BioLegend #79997). CD3 and CD19 positive viable cells were collected in BD FACS melody cell sorter using a 70 µm sort nozzle.

### V(D)J sequencing & analysis

10X Chromium platform BCR amplification and library preparation kits (10X Genomics) were used for preparing the libraries. Next generation sequencing was performed using Illumina NextSeq 550 platform. The BCR sequences for each single cell were assembled by the Cell Ranger pipeline (v5.0.0). After identification of the CDR3 sequences and the rearranged BCR genes, analysis was performed using Loupe V(D)J Browser v.2.0.1. BCR diversity metric, containing clonotype frequency and barcode information was processed using Seurat (v3) and plotted using GGPLOT2 R package.

### Neutralization assay

Neutralization assay was performed according to manufacturer’s protocol (SARS-CoV-2 Surrogate Virus Neutralization Test Kit, Genscript # L00847-A). Briefly, positive control, negative control and samples (serum 1:4 dilution) were diluted with HRP-RBD in a ratio of 1:1 in tubes and incubated at 37□ for 30 minutes. After incubation 100 µl of each mixture were added to the plate for 15 minutes at 37□. Following which the wells were washed with 1x wash solution 4x times and finally TMB substrate was added for development of colour. The reaction was stopped using 50 µL of stop solution and absorbance was measured immediately in a Multiskan reader (Thermo scientific). The percent inhibition/neutralization was calculated using the formula = (1 - OD value of Sample /OD value of Negative Control) × 100%.

### Statistical tests

All the statistical tests are performed in R and the bar and box plots are generated using the GGPUBR package. The pie chart of antibodies was created using the GGPLOT2 package and all the respective p-values in box plots are calculated using Wilcoxon test and stat_compare_means (paired = FALSE) function for respective condition pairs.

## Results

We characterized the antibody titre, SARS-CoV-2 surrogate virus neutralization efficacy, cytokine levels, and single cell V(D)J profiles of 61 individuals (18 females and 43 males) distributed among three categories (Supplementary Figure 1A-B, Supplementary Table1). We performed ELISA from serum to quantify individual antibodies specific to spike RBD. The overall antibody distribution of contact showed increased levels of IgG and IgA in comparison to others but the median levels of IgM were similar to symptomatic ones, although higher than control (Supplementary Figure 1C). Groupwise comparing mean the levels IgA, IgM and IgG antibody titres (Figure 1A), we observed contact individuals to have significant levels of IgA, IgM and IgG (p<0.05). Further quantification of antibody levels of symptomatic and asymptomatic individuals was done independently (Supplementary Figure 1B). Symptomatic patients showed elevated levels of IgA, IgM, and IgG in comparison to asymptomatic patients in the median level (the changes were not significant). Severe COVID-19 patients experience hyper-activation of immune responses, predominantly polyfunctionality in CD8^+^ T cells, distinct CD4^+^ T cell subpopulations and B-cell heterogeneity [9]. Similarly, the cytokine levels among mild, severe and fatal Covid-19 patients differs drastically [10]. For functional characterization, we determined the cytokine levels among the subject groups and found to be differentially secreted between contact and infected individuals. We observed that out of 48 cytokines 21 were able to distinguish between the two groups (data not shown) using logistic regression (Pr(>|z|) <0.05), among which 5 cytokines were showing prominent distinction viz. Eotaxin, G-CSF, IL-7, MIF and MIP1-α and their secretory levels are highly significant in contact with comparison to infected and control (Figure 1B). The area under the curve of the ROC (Receiver Operating Characteristic) line had more than 0.80 AUC (Figure 1C), suggesting an aggregated measure of performance between individuals. Further, to validate the COVID-19 neutralizing efficacy of the serum samples we performed an *in-vitro* spike RBD-ACE2 interaction based Surrogate Virus Neutralization assay for representative (Supplementary Table1) samples. Neutralizing antibodies against SARS-CoV-2 were detected in serum samples of contact, symptomatic and asymptomatic individuals in gradient (Figure 1D). The percent inhibition shown by contact individuals in accordance with control and infected are exceptionally noteworthy and correlate with high neutralizing antibody titers. For acquiring a holistic perspective of the clonotype changes associated with the BCR repertoires we performed single cell V(D)J sequencing and analysed BCR clonal expansion. We took a total of twelve samples for our study, unmatched control (n=3), asymptomatic (n=3), symptomatic (n=3) and contact (n=3). To understand the antibody response in patients, we plotted the distribution of IgA, IgM, IgG and IgD in each individual for each of the conditions respectively (Figure 2A). The cumulative percentage of IgA isotype increased in contact (depicted in individual CV0030 and CV0041) along with overall expansion of clonotypes compared to control and infected (Figure 2 A&B), suggesting that our result of high antibody production and clonotype expansion correlation might hold true. To study the gene preference of BCRs, we compared the differential usage of V(D)J genes across the conditions and observed over-representation of *IGHV4-59* and previously reported gene *IGHV3-15* [11]. The preferred IGKVs and IGLVs were *IGKV1D-39* and *IGLV1-51, IGLV2-11* respectively (Figure 2C and Supplementary Figure 1D) and also noticed over-representation of *IGHJ4* in all individual groups [12]. Overall increased BCR clonotypes and biased usage of genes in contacts can be the reason for high antibody production.

**Figure 1.**
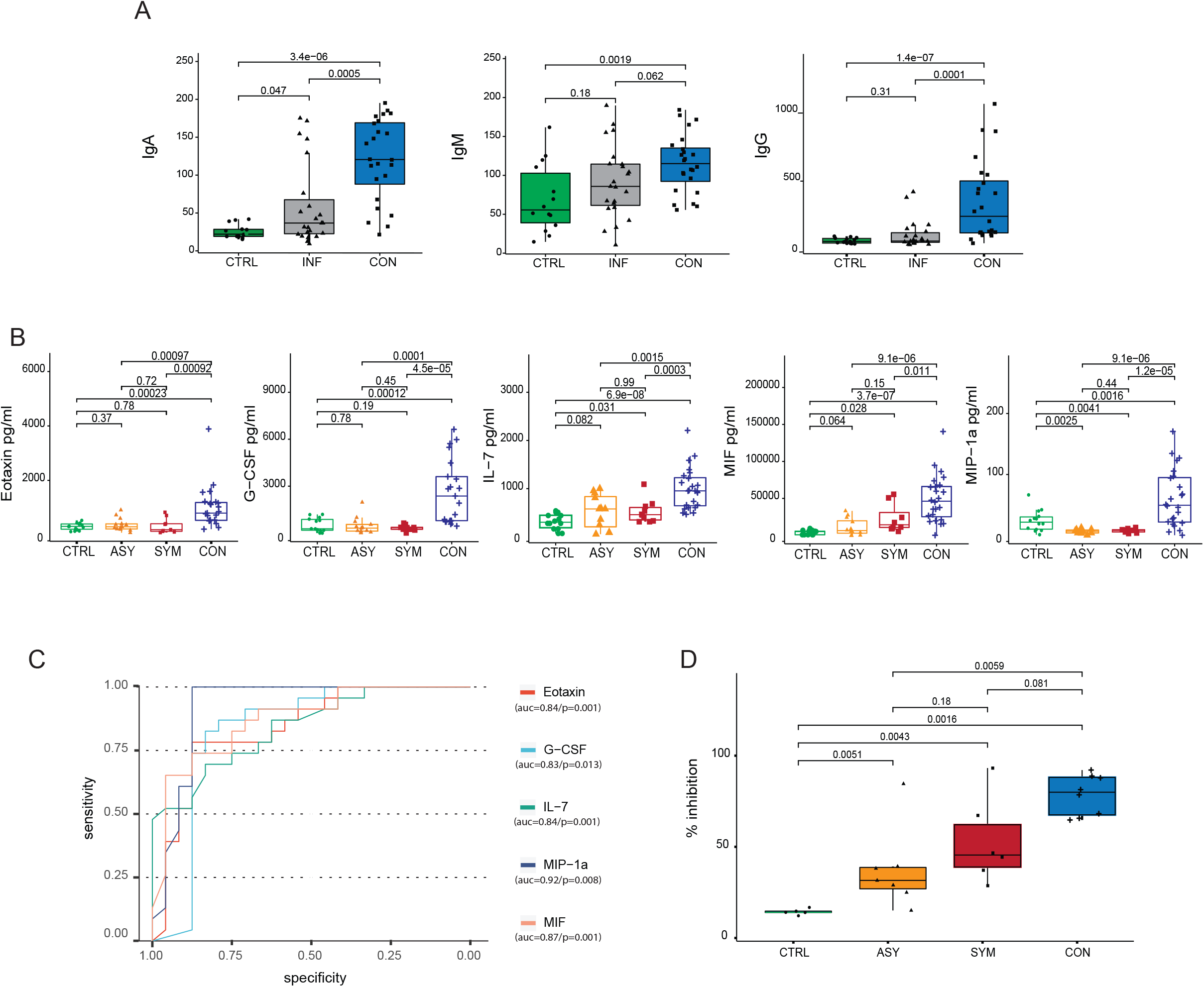
Immune response to SARS-CoV-2 antigen with neutralizing magnitude. **A**, Box plots showing the anti-SARS-CoV-2 IgA, IgM and IgG antibody proportions in serum samples from control, infected and contact along with **B**, Serum cytokine levels in pg/ml (Eotaxin, G-CSF, IL-7, MIF and MIP 1-□) determined by bio plex. **C**, The area under the receiver-operating characteristic (ROC) curve of the cytokines having more than 0.8 AUC. **D**, Box plot depicting the percent inhibition of neutralizing antibodies in an in*-vitro* spike RBD-ACE2 interaction based Surrogate Virus Neutralization assay. Statistical comparisons were performed using unpaired Wilcoxon test, p-values are written against respective comparisons. Where, n = number of individuals, IgA-Immunoglobulin A, IgM-Immunoglobulin M and IgG-Immunoglobulin G, control (n-14), infected (n-23, Symptomatic n-10 and Asymptomatic n-13) and contact (n-24).

**Figure 2.**
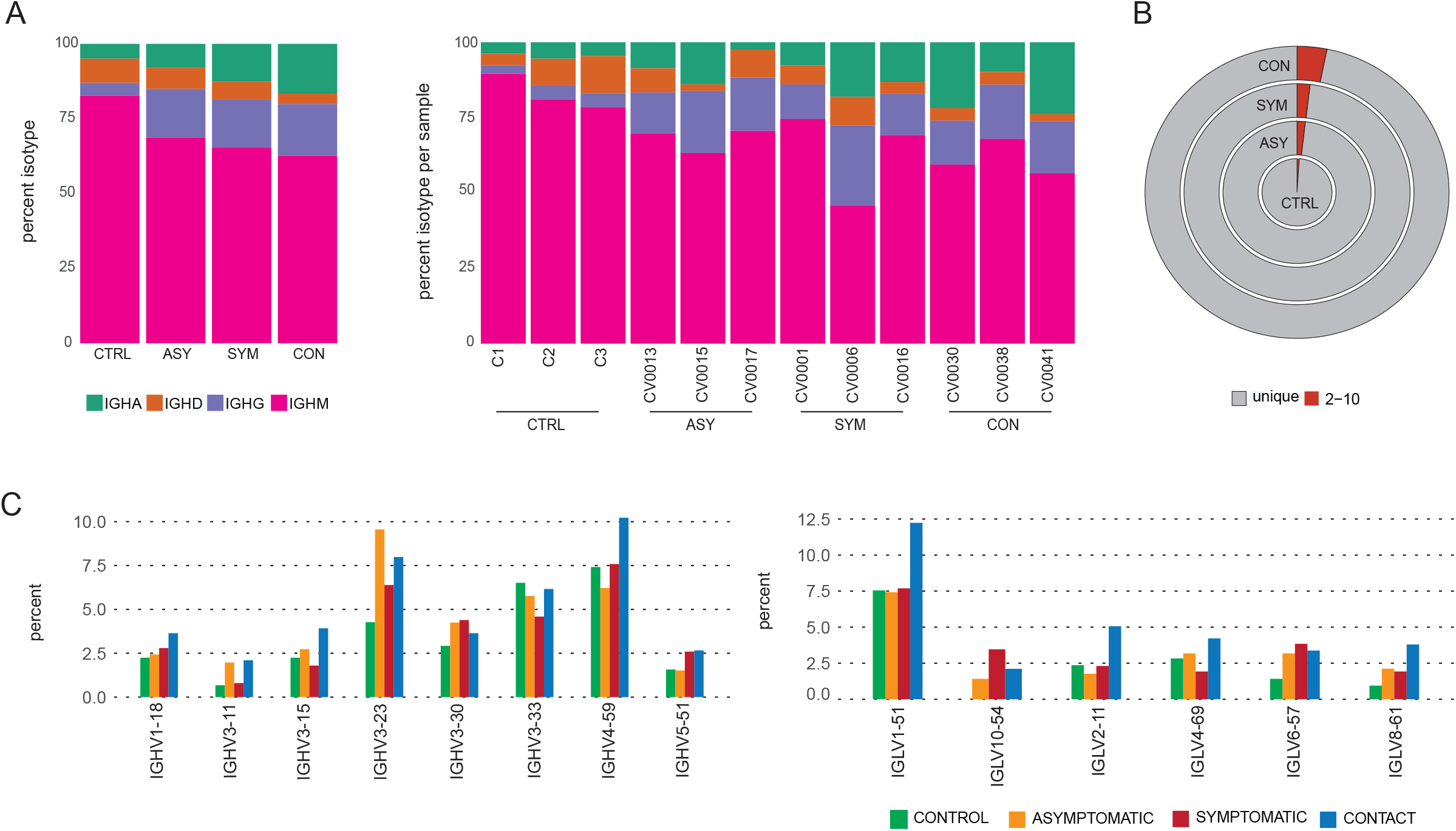
Distribution of IGs, BCR clonotypes and differential usage of V(D)J genes across groups. **A**, Bar plots showing percentages of IGHA, IGHG, IGHM and IGHD in the four conditions(left) and in each individual per condition(right). **B**, Multiple pie charts showing the percentage clonal status of B cells. **C**, Bar plots showing usage of some specific IGHV (lower-left) and IGLV (lower-right) genes across the four conditions.

## Discussion

The nature of protection observed in contact individuals is of great concern keeping in view the neutralizing efficacy towards SARS-CoV-2 antigen. The surged level of antibodies hints towards prior exposure with other pathogens having similar antigenic determinants. It is also exciting to note that when we correlated the antibody profile with the cytokine data, contacts showing elevated levels of antibodies were also showing high levels of particular cytokines. Eotaxin, G-CSF, IL-7 and MIP 1-α are secreted across all subject groups in higher quantities than MIP 1-α which is a signature cytokine for increased disease severity. Among which, IL-7 and G-CSF play pivotal roles in lymphocyte expansion and rendering protection against invading pathogens respectively. The inflammatory chemokine Eotaxin and pro-inflammatory macrophage derived factor MIF are also key players in mediating immune responses. So, co-occurrence of specific cytokines with elevated Ig levels appeared to be a protective signature among contacts. Specifically, IgA antibody is of keen interest since it has the capacity of neutralizing respiratory viruses and protecting mucosal surfaces by hindering their attachment to epithelial cells [6].

Direct contacts also showed specific BCR profiles and differential usage of genes along with unique clonotypes essential for high antibody production and cytokine secretion. All the above observations provided a strong and protective host-immune response as a major determining factor in contracting the COVID-19 disease.

## Data Availability

All data are available with the corresponding author and will be shared on request.

## Ethical approval and consent

The blood samples for this study were obtained from TATA COVID hospital, Ganjam District, Odisha after due approval of the Institutional Biosafety and human ethics committee (Ref: 101/HEC/2020). For collecting the blood samples for the study due consent was taken from the subjects involved in the study.

## Acknowledgments

We would like to thank all the volunteers who participated in the study voluntarily and provided their blood samples. We would also like to thank ILS core facilities (Single cell and genomics, Flow cytometry) and their staff for help and support. Also like to acknowledge ILS for institutional support for this study.

---

**Supplementary Figure 1. Gender and condition, antibody titre, distribution and biased usage of V(D)J genes across groups A-B** Bar graphs showing the distribution of male (n-43) and female (n-18) individuals and their categorization into different subject groups. **C**, Antibody levels in symptomatic and asymptomatic patients **C**, Pie-chart delineating the overall immunoglobulin distribution across subject groups and **D**, Bar plots showing usage of IGKV, IGKJ, IGHJ, and IGLJ genes across the four conditions. Conditions are shown in different colours. Statistical comparisons were performed using unpaired Wilcoxon test, p-values are written against respective comparisons.

