## Supplementary figures and images for "SARS-CoV-2 specific immune-signature in direct contacts of COVID-19 cases protect them from contracting disease: A Retrospective Study"

### Supplementary figure 1

#### Supplementary figure 1

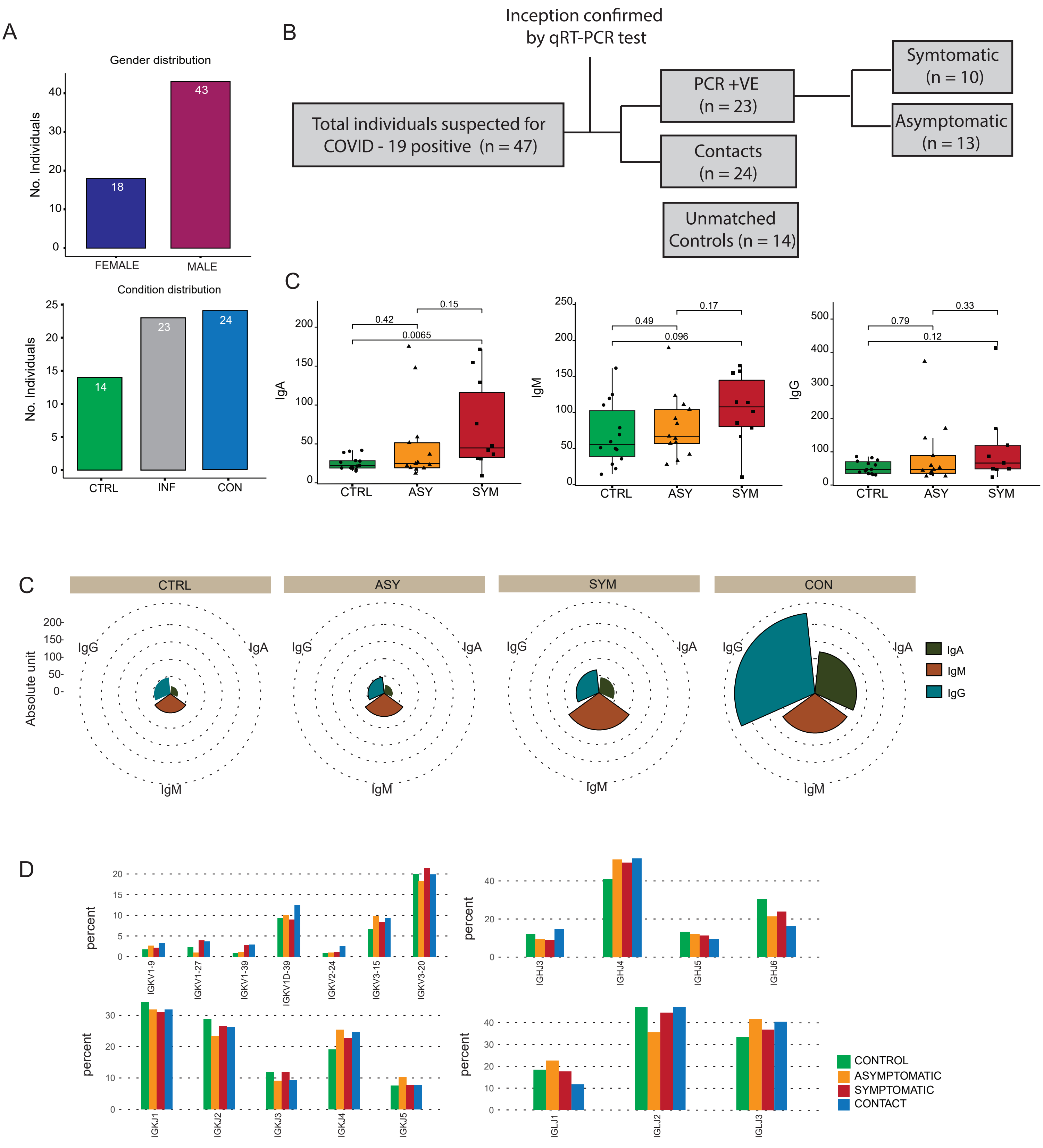
